# The dichotomous and incomplete adaptive immunity in COVID-19

**DOI:** 10.1101/2020.09.05.20187435

**Authors:** Leiqiong Gao, Jing Zhou, Sen Yang, Xiangyu Chen, Yang Yang, Ren Li, Zhiwei Pan, Jing Zhao, Zhirong Li, Qizhao Huang, Jianfang Tang, Li Hu, Pinghuang Liu, Guozhong Zhang, Yaokai Chen, Lilin Ye

## Abstract

The adaptive immunity that protects patients from coronavirus disease 2019 (COVID-19), caused by severe acute respiratory syndrome coronavirus 2 (SARS-CoV-2), is not well characterized. In particular, the asymptomatic patients have been found to induce weak and transient SARS-CoV-2 antibody responses, but the underlying mechanisms remain unknown; meanwhile, the protective immunity that guide the recovery of these asymptomatic patients is also not well studied. Here, we characterized SARS-CoV-2-specific B-cell and T-cell responses in 10 asymptomatic patients and 49 patients with other disease severity (mild, n = 10, moderate, n = 32, severe, n = 7) and found that asymptomatic or mild symptomatic patients failed to mount virus-specific germinal center (GC) B cell responses that result in robust and long-term humoral immunity, assessed by GC response indicators including follicular helper T (T_FH_) cell and memory B cell responses as well as serum CXCL13 levels. Alternatively, these patients mounted potent virus-specific T_H_1 and CD8^+^ T cell responses. In sharp contrast, patients of moderate or severe disease induced vigorous virus-specific GC B cell responses and associated T_FH_ responses; however, the virus-specific T_H_1 and CD8^+^ T cells were minimally induced in these patients. These results therefore uncovered the protective immunity in COVID-19 patients and revealed the strikingly dichotomous and incomplete adaptive immunity in COVID-19 patients with different disease severity, providing important insights into rational design of COVID-19 vaccines.

## Introduction

As of July 19, 2020, the ongoing pandemic of COVID-19, caused by SARS-CoV-2 infection, has led to over 14 million confirmed cases and 597 thousand deaths, according to WHO issued COVID-19 Situation Report-181(WHO, 2020). Thus far, no vaccines has been approved to prevent SARS-CoV-2 infection, albeit several types of vaccine candidates reported at different clinical trial stages (Amanat and Krammer, 2020; Thanh Le et al., 2020).

The SARS-CoV-2 infected patients generally manifest diverse clinical symptoms, ranging from no symptoms to critical illness, which can be further categorized into four groups, including asymptomatic, mild, moderate and severe(2020; Mizumoto et al., 2020; Raoult et al., 2020). The adaptive immunity, encompassing humoral and cellular immune responses, is a key to clear a wide variety of viral infections, rendering patients recovered from viral diseases (Aoshi et al., 2011). In SARS-CoV-2 infection, both virus-specific B-cell mediated humor immunity and T-cell mediated cellular immunity have been implicated in recovered COVID-19 patients(Cao et al., 2020; Grifoni et al., 2020; Le Bert et al., 2020; Ni et al., 2020; Woodruff et al., 2020). Notably, in asymptomatic patients, SARS-CoV-2-specific IgGs were minimally produced and poorly maintained (Ibarrondo FJ, 2020), in contrast, patients of severe disease mounted potent virus-specific IgG responses(Lee et al., 2020; Long et al., 2020; Zhang et al., 2020). However, the immune mechanisms underlying the dichotomous virus-specific IgG immune responses between asymptomatic or severe symptomatic patients are not well understood. Thus far, it also remains unknown whether virus-specific T cell immune responses are effectively induced to protect asymptomatic patients from progressing to severe disease. To characterize virus-specific B- and T- cell immune responses in recovered COVID-19 patients with different degrees of clinical symptoms will provide important insights into understanding the protective immunity for COVID-19, which will lay the foundation for rationally designing effective vaccines against SARS-CoV-2 infection.

## Results

### subjects

To explore the adaptive immune responses in recovered COVID-19 patients with different clinical symptoms, we organized a cohort of 59 adult patients (29 males and 30 females), including asymptomatic (n = 10), mild (n = 10), moderate (n = 32), severe (n = 7) symptoms, admitted to Chongqing Public Health Center, China (Table S1&S2). All the patients were positive with SARS-CoV-2 nucleic acid testing. The clinical and pathological characteristics were summarized in Table S2. The disease severity was stratified into asymptomatic, mild, moderate and severe based on the national diagnosis and treatment guideline of COVID-19 (7^th^ edition) in China(2020) (Table S1). The admission date was from 02/06/2020 to 04/24/2020 and the average duration of hospitalization was 19 (7-51) days. The peripheral blood mononuclear cells (PBMCs) and sera were collected at 10 to 17-day post-discharge for all the patients. For asymptomatic patients, their PBMCs were harvested twice during the hospitalization.

### Failure of SARS-CoV-2-specific GC B cell reaction in asymptomatic COVID-19 patients

By flow cytometry analysis, we found comparable proportions of CD4^+^, CD8^+^ T cells and B cells among PBMCs between patients with asymptomatic, mild illness and healthy controls. However, moderate, and severe symptomatic patients exhibited a significant reduction in proportions of CD4^+^ and CD8^+^ T cells but a marked increase in frequencies of B-cells compared to healthy controls (Figure. S1A–S1C), consistent with the observations in recent studies (Wen et al., 2020; Woodruff et al., 2020).

The increase of total B cell compartment in recovered COVID-19 patients of moderate and severe illness likely indicated elevated SARS-CoV2-specific B cell responses. To test this hypothesis, we investigated B cells specifically recognizing SARS-CoV-2 spike protein subunits, S1 and S2, both of which represent dominant antigens of SARS-CoV-2 to induce virus-specific B-cell responses(Chen et al., 2020; Ho, 2020). As expected, we noted remarkably increased S1/S2-specific B cells in recovered patients with moderate or severe symptoms compared to those in recovered patients of asymptomatic or mild illness (Figure. 1A, 1C). Coincident with these results, we also observed a substantial enhanced S1/S2-specific memory B cell population (IgD^low^CD19^+^S1^+^/S2^+^CD27^hi^) in patients with moderate (about 10% of S1/S2-specific B cells) or severe disease (about 25% of S1/S2-specific B cells), while memory B-cell fractions were scarcely shown in patients of asymptomatic or mild illness (Figure.1B, 1D). In line with these results, we found S1/S2-specific IgG titers highest in sera of patients with moderate or severe disease, while lowest in sera of patients with asymptomatic or mild disease, consistent with the results observed in recent studies (Long et al., 2020) (Figure.1E, 1F).

**Figure.1.**
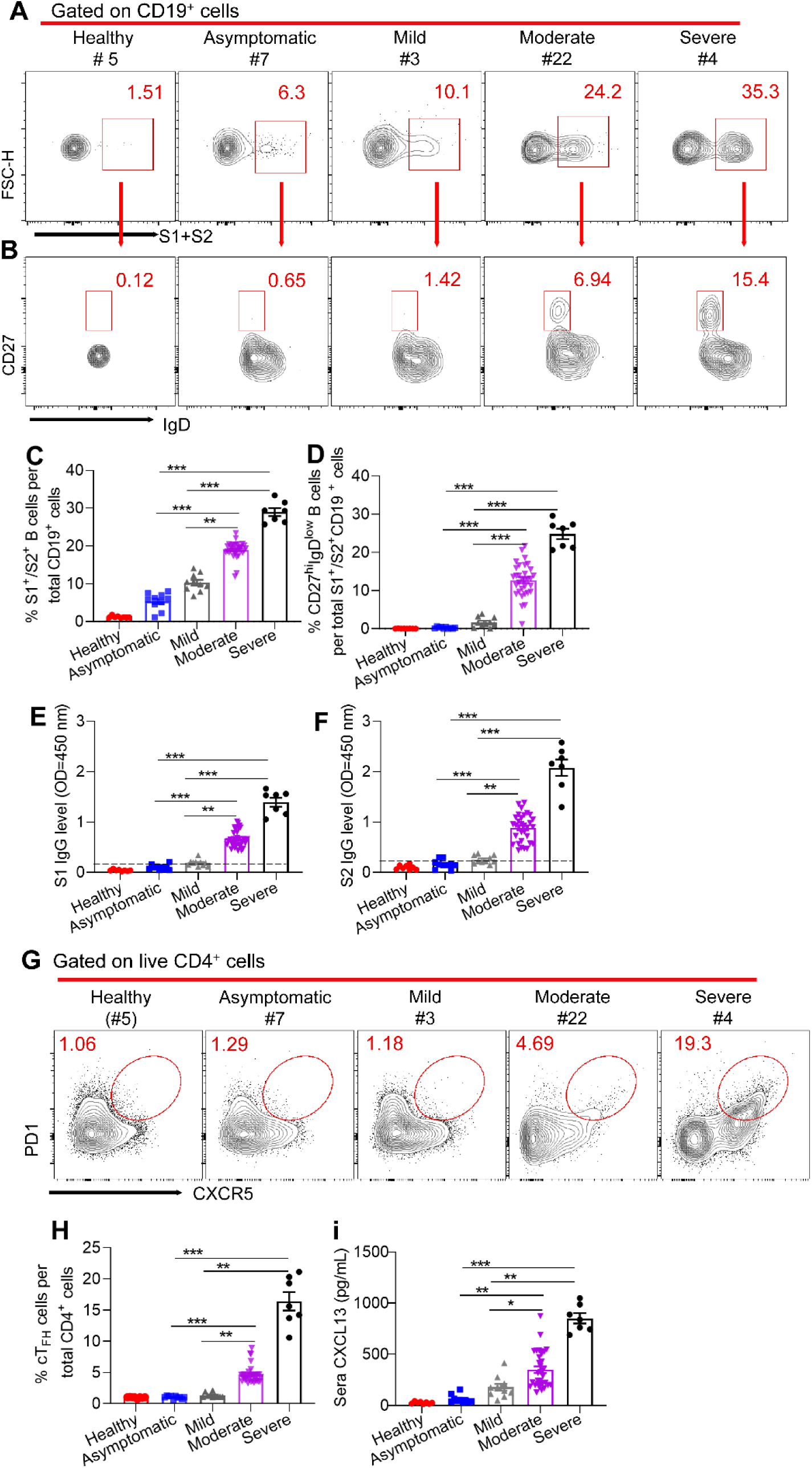
Virus-specific B cell responses to SARS-CoV2 in COVID-19 patients with different severity. Samples of **A, B** and **G** were from Healthy (#5), Asymptomatic (#7), Mild (#3), Moderate (#22), Severe (#4). **A**, FACS plot examples of SARS-CoV2 S1- or S2- specific B cells (S1^+^/S2^+^CD19^+^) of live lymphocytes in PBMCs. **B**, FACS plot examples of SARS-CoV2 S1- or S2- specific memory B cells (CD27^hi^IgD^low^), gated on S1 or S2 specific B cells (S1^+^/S2^+^CD19^+^) shown in **A. C & D**, Percentages of S1 or S2 specific B cells (**C**), summarized from (**A**); and S1 or S2 specific memory B cells (**D**), summarized from (**B**), in COVID-19 recovered patients with different disease severity. **E & F**, ELISA analysis of S1- and S2- specific IgG levels in convalescent-phase COVID-19 patients’ sera with various groups. Dilution of 1:100 was used for serum samples. **G**, FACS plot examples of cT_FH_ (PD1^hi^CXCR5^hi^), gated on live CD4^+^ T cells. **H**, The summarization of percentages of cT_FH_ from (**G**). **I**, CXCL13 protein level in convalescent-phase COVID-19 patients’ serum with various groups, measured by ELISA. Bars represent the mean ± SEM. *P* values were calculated based on Bonferroni of one-way analysis. ***, *p*<0.0001, **, *p*<0.001, and *, *p*<0.05.

Germinal center (GC) reaction in B-cell follicles within secondary lymphoid tissues gives rise to long-lived memory B cells and bone-marrow resident plasma cells capable of constitutively secreting antigen-specific IgGs, which strictly depends on the help provided by cognate follicular helper T (T_FH_) cells(Huang et al., 2019; Pedros et al., 2016). Since lymphoid tissues are inaccessible from recovered COVID-19 patients, we took advantage of measuring circulating T_FH_ (cT_FH_) cell and chemokine CXCL13 to reliably indicate the magnitude of ongoing GC responses(Brenna et al., 2020; McGuire et al., 2019). In support of aforementioned results, we noted a PD1^hi^CXCR5^hi^ cT_FH_ population accounting for about 15% of total CD4^+^ T cells in COVID-19 patients recovered from severe disease, about 5% in patients of moderate disease, while such population was almost unappreciable in counterparts with asymptomatic or mild disease (Figure. 1G, 1H). Likewise, we observed the minimal CXCL13 concentration in sera from recovered patients with asymptomatic or mild illness; In stark contrast, copious CXCL13 was detected in sera from patients recovered from moderate (average 415 pg/ml) or severe disease (average 850 pg/ml) (Figure. 1I). Further correlation analysis revealed the high relevance between S1/S2-specific IgG antibodies and total B-cell frequencies, CXCL13 concentration and frequencies of cT_FH_ (Figure. S2A-S2F). We also observed the high correlation between CXCL13 concentration and frequencies of cT_FH_ (Figure. S2G).

### Transient SARS-CoV-2-specific B cell response in asymptomatic individuals

Given the failure of inducing prolonged SARS-CoV-2-specific B-cell responses by asymptomatic or mild patients, we wonder whether these patients have mounted transient virus-specific B-cell responses during their hospitalization. To this end, we first analyzed S1- or S2-specific B cell frequencies in their PBMCs at different time points during hospitalization. We found that the peak of S1/S2-specific B cell responses was induced at early stage (day 0-3) of hospitalization, which rapidly waned at the middle and convalescent stages (Figure. 2A, 2C). Concomitantly, we observed the similar dynamics of plasmablasts (CD19^+^CD20^−^CD27^hi^CD38^hi^) (Figure.2B, 2D), which generally reflect the extrafollicular antibody responses during primary viral infection(Carter et al., 2017; Fink, 2012; Woodruff et al., 2020). In keeping with these observations, we also noticed the relatively higher S1- and S2- specific IgG titers at the middle stage (5-10 day post hospitalization) of hospitalization than those at both early (0-3 day post hospitalization) and convalescent stages (more than 10 days after COVID-19 nucleic acid test shown negative) (Figure. 2E). Furthermore, we found nearly no cT_FH_ population induced and CXCL13 secreted from early phase of hospitalization to convalescent stages (Figure. 2F-2H). These data collectively revealed that SARS-CoV-2-specfiic B cell responses were only transiently induced in asymptomatic patients and sustained GC responses that give rise to long-term memory B cells and IgG-secreting plasma cells were likely absent in these patients, explaining the weak and short-term SARS-CoV-2 specific IgGs in these patients reported most recently (Long et al., 2020). Taken together, our results demonstrate that compared to potent and sustained SARS-CoV-2-specific GC B cell responses mounted in COVID-19 patients recovered from moderate or severe symptoms, asymptomatic or mild symptomatic COVID-19 patients only induced weak and transient SARS-CoV-2-specific B cell responses.

**Figure.2.**
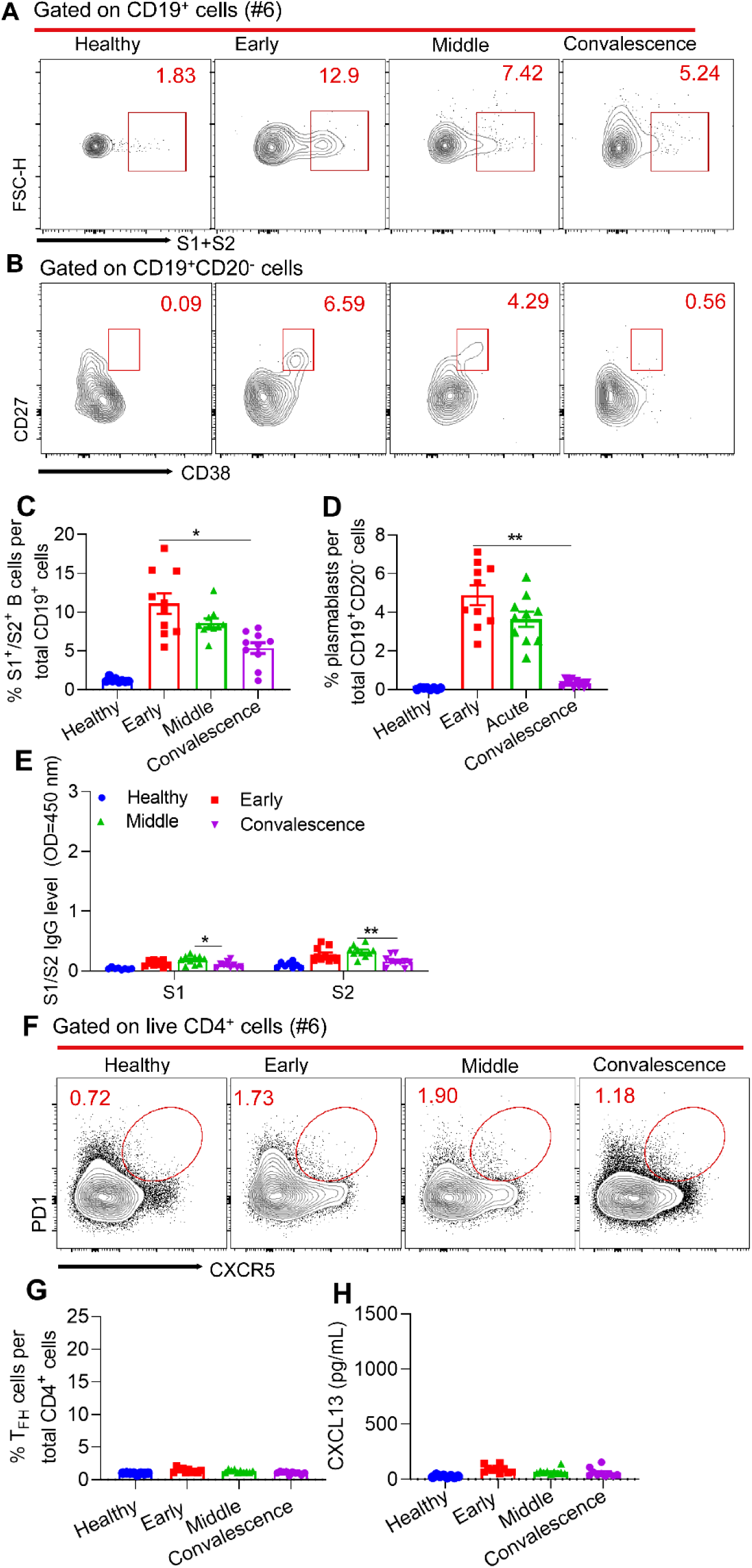
Kinetic virus-specific B cell responses to SARS-CoV2 in COVID-19 Asymptomatic patients. Sample of **A, B** and **F** was from Asymptomatic (#6). **A & B**, FACS plot examples of SARS-CoV2 S1- or S2- specific B cells and plasmablasts (CD19^+^CD20^−^CD27^hi^CD38^hi^) cells’ percentage in early phase (0-3 day post hospitalization), acute phase (5-10 day post hospitalization), convalescent-phase, (more than 10 days after COVID-19 nucleic acid test shown negative) in the asymptomatic cohort. **C & D**, Percentages of S1- or S2- specific B cells (**C**), summarized from (**B**), and plasmablasts cells (**D**), summarized from (**B**), in the asymptomatic cohort. **E**, Kinetics of S1- and S2- specific IgG levels in asymptomatic patients’ serum, measured by ELISA. **F**, FACS plot examples of cT_FH_ cells’ percentage of asymptomatic patients. **G**, Percentages of cT_FH_, summarized from (**O)**. **H**, Kinetics of CXCL13 level in asymptomatic patients’ serum, measured by ELISA. Bars represent the mean ± SEM. *P* values were calculated based on Bonferroni of one-way analysis. ***, *p*<0.0001, **, *p*<0.001, and *, *p*<0.05.

### Potent SARS-CoV-2 specific cellular immune responses in COVID-19 patients with asymptomatic disease

In addition to B-cell associated humoral immunity, the cellular immunity mediated by T_H_1 and cytotoxic CD8^+^ T lymphocytes also plays a critical role in the control of viral infection(Aoshi et al., 2011; Chen et al., 2005; Janice Oh et al., 2012; Mahallawi et al., 2018; Wong et al., 2004; Yoo et al., 2013). The failure in the induction of a protective humoral immunity by COVID-19 patients recovered from asymptomatic or mild disease may indicate an alternative strong cellular immunity that protected these patients from developing severe disease. To test this hypothesis, we sought to examine SARS-CoV-2-specific cellular immunity in recovered COVID-19 patients of different disease severity. To define the SARS-CoV2-specific CD4^+^ and CD8^+^ T cells, we stimulated total PBMCs with SARS-CoV-2 dominant antigen (S1, S2 and nucleoprotein, N) cocktails for 48 hours(Ni et al., 2020). After antigen stimulation, we observed the background levels of IFN-γ producing CD4^+^ and CD8^+^ T cells in PBMCs of healthy controls; in contrast, in asymptomatic patients, the proportions of IFN-γ producing CD4^+^ and CD8^+^ T cells were substantially enhanced (Figure. 3A, 3B, 3E, 3F), indicative of the specificity of our strategy in analyzing SARS-CoV-2-specific CD4^+^ and CD8^+^ T cells. Next, we compared the proportions of T_H_1 cells that secrete hallmark cytokine IFN-γ in PBMCs among patients of different disease severity. Strikingly, we found the remarkably upregulated abundances of virus-specific IFN-γ-secreting T_H1_ cells in asymptomatic or mild symptomatic patients relative to healthy controls; however, in patients with moderate or severe symptoms, they only exhibited background levels of virus-specific IFN-γ-producing T_H1_ cells post antigen stimulation as healthy control (Figure. 3A, 3B). Moreover, among those IFN-γ-producing T_H_1 cells, around 10% of them were found to express cytolytic molecules granzyme B (GZMB) and perforin in patients of asymptomatic or mild illness, both of which have been found to be critical for the control of respiratory viral infection(Takeuchi and Saito, 2017), while only around 2.5% of GZMB and perforin-producing T_H_1 cells in patients with moderate or severe disease (Figure. 3C, 3D). These results together indicated that virus-specific CD4^+^ T cell responses was much biased toward to T_H_1 over T_FH_ cells in asymptomatic or mild symptomatic patients, while vice versa in patients with moderate or severe disease. The cellular and molecular mechanisms underlying disease severity-dependent virus-specific CD4^+^ T-cell differentiation in COVID-19 patients warrants further investigations.

**Figure.3.**
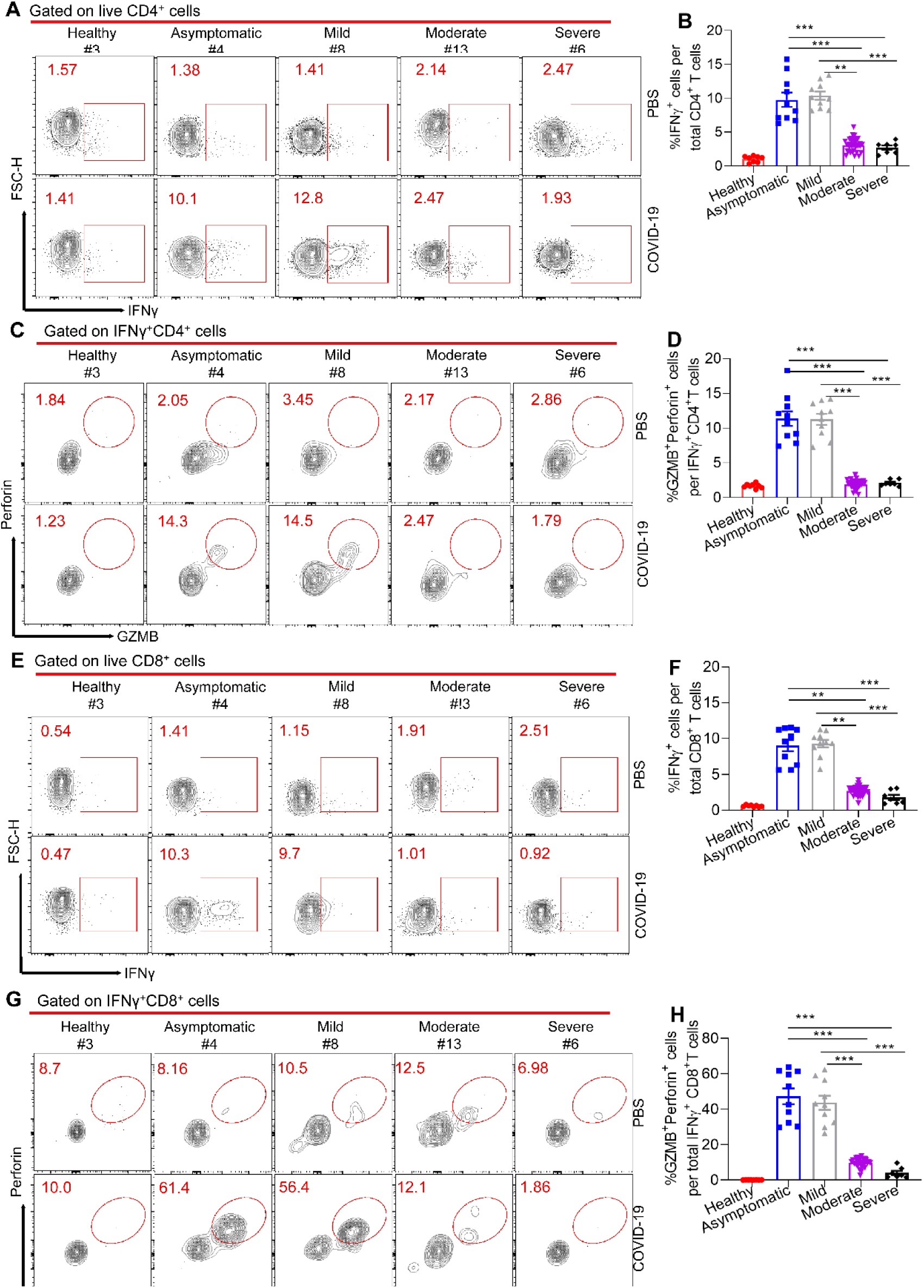
SARS-CoV2 specific CD4^+^ and CD8^+^ T cell responses in COVID-19 convalescent patients. PBMCs of healthy control (n = 8) and recovered COVID-19 patients (n = 59) were stimulated with SARS-CoV-2 dominant antigen (S1, S2 and nucleoprotein, N) cocktails for 44 hours, Golgi-Plug containing Golgi-stop and DNAase were added into cell culture for another 4 hours. Samples of **A, C, E** and **G** were from Healthy (#3), Asymptomatic (#4), Mild (#8), Moderate (#13), severe (#6). **A**, FACS plot examples of IFNγ^+^CD4^+^ T cells in total live CD4^+^ T cells, gated on total live CD4^+^ T cells. **B**, Bar graph shows frequency of IFNγ^+^CD4^+^ T cells in total CD4^+^ T cells after stimulation. **C**, FACS plot examples of GZMB^+^Perforin^+^CD4^+^ T cells in total IFNγ^+^CD4^+^ T cells, gated on total live IFNγ^+^CD4^+^ T cells. **D**, Frequency of GZMB^+^Perforin^+^CD4^+^ T cells in total IFNγ^+^CD4^+^ T cells, summarized form (**C**). **E**, FACS plot examples of IFNγ^+^CD8^+^ T cells in total live CD8^+^ T cells, gated on total live CD8+ T cells. **F**, Bar graph shows frequency of IFNγ^+^CD8^+^ T cells in total CD8^+^ T cells after stimulation. **G**, FACS plot examples of GZMB^+^Perforin^+^CD8^+^ T cells in total IFNγ^+^CD8^+^ T cells, gated on total live IFNγ^+^CD8^+^ T cells. **H**, Frequency of GZMB^+^Perforin^+^CD8^+^ T cells in total IFNγ^+^CD8^+^ T cells, summarized form (**G**). Bars represent the mean ± SEM. *P* values were calculated based on Bonferroni of one-way analysis. ***, *p*<0.0001, **, *p*<0.001, and *, *p*<0.05.

Next, we assessed SARS-CoV-2-specific CD8^+^ T cell responses in COVID-19 recovered patients. Similar to the results of virus-specific T_H_1 responses, we found substantially induced virus-specific IFN-γ-producing CD8^+^ T cells in asymptomatic or mild symptomatic patients as compared to healthy controls, whereas very limited IFN-γ-producing CD8^+^ T cells in patients of moderate or severe symptoms (Figure. 3E, 3F). To further assess the cytolytic functionality of virus-specific CD8^+^ T cells in COVID-19 patients, we first gated on virus-specific IFN-γ^+^ CD8^+^ T cells, followed by measuring the frequencies of cells capable of producing GZMB and perforin. We found that in asymptomatic or mild symptomatic patients, approximate 60% of IFN-γ^+^ CD8^+^ T cells simultaneously produced GZMB and perforin, whereas only 12% or 3% of IFN-γ^+^ CD8^+^ T cells were able to make both cytolytic molecules in moderate or severe patients (Figure.3G, 3H). These data therefore revealed that albeit the failure in mounting SARS-CoV-2-specific humoral immunity, asymptomatic or mild symptomatic patients were able to induce a profound virus-specific cellular (T_H1_ and CD8^+^ T cell) immunity. In sharp contrast, patients of moderate or severe disease were able to generate potent SARS-CoV-2-specific humoral immunity, but they failed to induce effective virus-specific cellular immunity. The mechanisms underlying disease severity-associated dichotomous humoral and cellular immunity in COVID-19 patients need to be investigated in the future.

### Early induced SARS-CoV-2 specific cellular immune responses in asymptomatic or mild individuals

Given the vigorous cellular responses detected in recovered COVID-19 patients of asymptomatic or mild disease, we wonder whether these patients have induced strong good virus-specific T-cell responses at the early phase during their hospitalization. After SARS-CoV-2 antigen stimulation, we found that IFN-γ producing CD4^+^ and CD8^+^ T cells was increased at early stage (day 0-3) of hospitalization in these asymptomatic patients, which were then well maintained at the middle (day 5-10) of hospitalization and convalescent stages (Figure. 4A, 4B, 4E, 4F). Concomitantly, we observed the similar dynamics of frequencies of T cells capable of producing GZMB and perforin in these asymptomatic patients (Figure.4C, 4D, 4G, 4H). These data collectively revealed that SARS-CoV-2-specfiic T_H_1 and CD8^+^ T cell responses were rapidly induced and sustained in asymptomatic patients.

**Figure.4.**
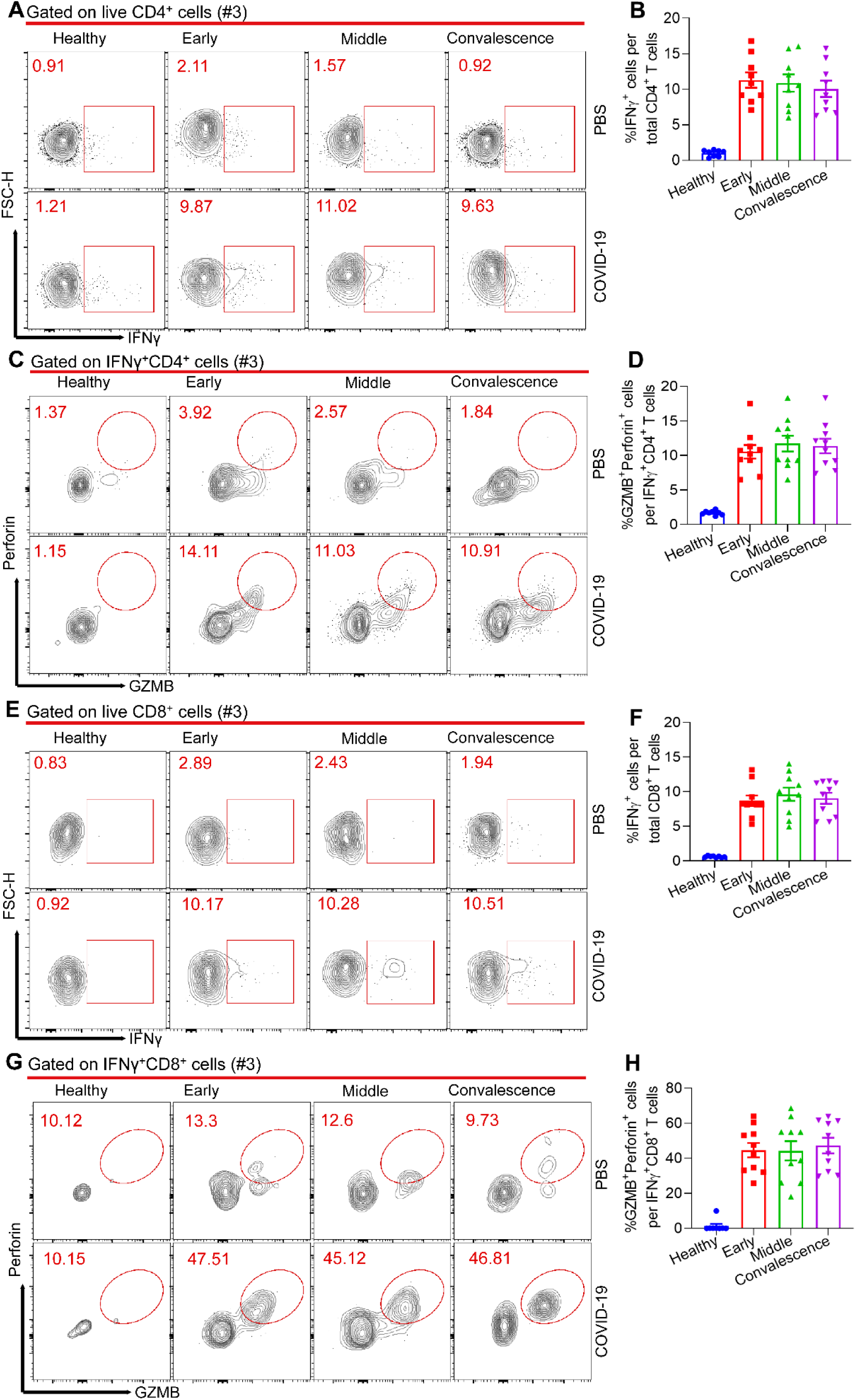
Kinetic SARS-CoV2 specific CD4^+^ and CD8^+^ T cell responses in COVID-19 asymptomatic patients. PBMCs of healthy control (n = 8) and asymptomaticCOVID-19 patients (n = 10) were stimulated with SARS-CoV-2 dominant antigen (S1, S2 and nucleoprotein, N) cocktails for 44 hours, Golgi-Plug containing Golgi-stop and DNAase were added into cell culture for another 4 hours. Representative samples were from Healthy (#3), Asymptomatic (#3). **A**, FACS plot examples of IFNγ^+^CD4^+^ T cells in total live CD4^+^ T cells, gated on total live CD4^+^ T cells. **B**, Bar graph shows frequency of IFNγ^+^CD4^+^ T cells in total CD4^+^ T cells after stimulation. **C**, FACS plot examples of GZMB^+^Perforin^+^CD4^+^ T cells in total IFNγ^+^CD4^+^ T cells, gated on total live IFNγ^+^CD4^+^ T cells. **D**, Frequency of GZMB^+^Perforin^+^CD4^+^ T cells in total IFNγ^+^CD4^+^ T cells, summarized form (**C**). **E**, FACS plot examples of IFNγ^+^CD8^+^ T cells in total live CD8^+^ T cells, gated on total live CD8+ T cells. **F**, Bar graph shows frequency of IFNγ^+^CD8^+^ T cells in total CD8^+^ T cells after stimulation. **G**, FACS plot examples of GZMB^+^Perforin^+^CD8^+^ T cells in total IFNγ^+^CD8^+^ T cells, gated on total live IFNγ^+^CD8^+^ T cells. **H**, Frequency of GZMB^+^Perforin^+^CD8^+^ T cells in total IFNγ^+^CD8^+^ T cells, summarized form (**G**). Bars represent the mean ± SEM. *P* values were calculated based on Bonferroni of one-way analysis. There was no statistically significant difference among different stage of asymptomatic patients.

## Discussion

Similar to most recent reports (Ibarrondo FJ, 2020; Long et al., 2020) our study found that anti-SARS-CoV-2-antibodies were rapidly decay in asymptomatic or mild patients. We further found SARS-CoV-2-specfic B cell responses were only transiently induced in early infection phase in asymptomatic or mild patients. In keeping with these observations, sustained GC responses that give rise to long-term memory B cells and IgG-secreting plasma cells were almost absent in these patients. These results therefore explained the phenomenon that asymptomatic patients failed to generate and maintain a long-term SARS-CoV-2-specific IgG response.

In contrast to humoral immunity, we found that the virus-specific T_H_1 and CD8^+^ T cell immune responses were rapidly induced and sustained in asymptomatic or mild symptomatic patients as compared to patients with moderate or severe disease, which presumably protect them from progressing to severe COVID-19. We also envision that the rapid and robust virus-specific T_H_1 and CD8^+^ T cell responses may effectively curtail the SARS-CoV-2 replication, which results in the inefficient viral antigen production and limits the GC reaction that requires sufficient antigen stimulation. Memory T cells induced by previous pathogens can protect the individual from re-infecting the similar pathogens with common epitopes and shape the clinical severity of subsequent infections (Welsh and Selin, 2002). Besides SARS-CoV-1 and MERS-CoV, there are another four virus, which are endemically transmitted and cause the common cold (OC43, HKU1, 229E and NL63) (Cui et al., 2019). Recently studies found there exists cross-reactive T cell recognition between circulating ‘common cold’ coronaviruses and SARS-CoV-2(Grifoni et al., 2020; Le Bert et al., 2020). It is therefore of great interest to examine whether the history of “common cold” coronavirus infection with pre-existing SARS-CoV-2 cross-reactive T cells could determine the clinically asymptomatic state in COVID-19 patients.

In summary, we revealed a striking dichotomous pattern of humoral and cellular immunity induced in patients of asymptomatic/mild or moderate/severe disease. The highly induced virus-specific T_H_1 and CD8^+^ T cell immune responses in asymptomatic or mild symptomatic patients may protect them from progressing to severe COVID-19 in the absence of humoral immunity, while potent virus-specific B cell responses likely account for the recovery of patients of moderate or severe COVID-19. These results supported the notion that SARS-CoV-2 infection generally induced paralyzed adaptive immunity, either humoral or cellular, suggesting that the induction of both optimal humoral and cellular immunity may be crucial for an effective prophylactic vaccine to prevent SARS-CoV-2 infection.

## Data Availability

The data that support the findings of this study are openly available in medRxiv

## Acknowledgement

This work was supported by grants from the National Science and Technology Major Project (No. 2017ZX10202102-006-002 to L.Y.), the National Natural Science Fund for Distinguished Young Scholars (No. 31825011 to L.Y.), the National Key Research Development Plan (No.2016YFA0502202 to L. Ye) and the Chongqing Special Research Project for Novel Coronavirus Pneumonia Prevention and Control (No. cstc2020jscx-2 to L.Y.; No. cstc2020jscx-fyzx0074 to Y.C.; ocstc2020jscx-fyzx0135 to Y.C.).

## Author contributions

L.G., J.Z., S.Y., X.C., Y.Y., R.L., Z.P., J.Z., Z.L., Q.H., J.T., and L.H. performed the experiments.

L.Y. designed the study, analyzed the data and wrote the paper with L.G., J.Z., S.Y., P.L., G.Z. and Y.C.; and L.Y., G.Z. and Y.C. supervised the study.

## Conflict of interest

The authors declare no competing interests.

## Material and Methods

### Human subjects

Blood samples from 8 healthy adult donors were obtained by the Institute of Immunology of Army Medical University, which has no contact with SARS-CoV-2 and test negative for SARS-CoV-2 RNA. These donors had no known history of any significant systemic diseases, including but not limited to, for example, autoimmune disease, kidney or liver disease, congestive heart failure, malignancy, coagulopathy, hepatitis B or C, or HIV infection.

The 59 COVID-19 convalescent donors enrolled in the study were provided written informed consent. The blood samples of COVID-19 patients were obtained from Chongqing Public Health Medical Center. The study received IRB approvals at Chongqing Public Health Medical Center (2020-023-01-KY).

### SARS-CoV-2 S1, S2 ELISA

ELISA protocol generally followed that of precious study(Chen et al., 2020). Briefly, costar 96-well clear plates (Costar, 42592) were coated with 500ng/mL SARS-CoV-2 S1 protein (Sino Biological, 40591-V08H) or SARS-CoV-2 S2 protein (Sino Biological, 40590-V08B) overnight at 4°C. The next day, plates were blocked with 100 μL blocking buffer (5% FBS and 0.1% Tween 20 in PBS) at room temperature for 2 hours. After washing with PBST buffer (0.1% Tween 20 in PBS), 1:100 diluted serum was then added to the plates and incubated for 1 hour at room temperature. Serum was diluted in blocking buffer. Serum was heat inactivated at 56°C for 30 minutes before added to the plate. Then, these plates were washed 5 times with 0.05% PBS-Tween 20. Then these ELISA plates were incubated with anti-human IgG HRP antibody (Bioss Biotech, 0297D) at room temperature for 1 hour. Anti-human IgG antibody was used at a 1:3000 dilution. Then, these plates were washed 5 times with 0.05% PBS-Tween 20 and 100 μL TMB buffer (Beyotime, P0209) was added and reacted for 15 minutes at room temperature. These reactions were stopped with 1M H_2_SO_4_ stopping buffer. Plates were read on a Beckman Coulter Plate Reader at 450 nm, and ODs were background subtracted.

### CXCL13 ELISA

The costar 96-well clear plates (Costar, 42592) were coated with 2μg/mL CXCL13 monoclonal antibody (Sino Biological, 70057-MM13) overnight at 4°C. The next day, plates were blocked with 100 μL blocking buffer (5% FBS and 0.1% Tween 20 in PBS) at room temperature for 2 hours. After washing with PBST buffer (0.1% Tween 20 in PBS), heat-inactivation serum was then added to the plates and incubated for 2 hours at room temperature. Then, these plates were washed 5 times with 0.05% PBS-Tween 20. Then these ELISA plates were incubated with 2μg/mL CXCL13 polyclonal antibody (Sino Biological, 70057-RP01) for 2 hours at room temperature. These plates were washed 5 times with 0.05% PBS-Tween 20. Then incubated with goat anti-rabbit IgG HRP antibody (Sigma) at room temperature for 1 hour. Anti-rabbit IgG antibody was used at a 1:3000 dilution. Then, these plates were washed 5 times with 0.05% PBS Tween 20 and 100 μL TMB buffer (Beyotime, P0209) was added and reacted for 15 minutes at room temperature. These reactions were stopped with 1M H_2_SO_4_ stopping buffer. Plates were read on a Beckman Coulter Plate Reader at 450 nm, and ODs were background subtracted.

### PBMC isolation and serum collection

Whole blood in micro-anticoagulant tube was centrifuged for 15 min at 2200 rpm at room temperature to separate the cellular fraction. Peripheral blood mononuclear cells (PBMCs) were isolated by density-gradient sedimentation using Ficoll-Paque(Haoyang Biological, Tianjin, China). Briefly speaking, blood 1:1 diluted with PBS, was gently layered over an equal volume of Ficoll in a 15 ml BD centrifuge tube and centrifuged for 25 minutes at 2200rpm without brake. There were four layers, the second layer contained PBMCs. These cells could be gently removed using a Pasteur pipette and added to PBS to wash off any remaining platelets. Isolated PBMCs were cryopreserved in cell recovery media (90% heat inactivated fetal bovine serum and 10% DMSO, Gibco) and stored in liquid nitrogen until used in the assays. The serum was carefully removed from blood without sodium citrate treatment and stored at −80°C.

### SARS-CoV2 recombinant protein S1/S2 biotinylation

For biotinylation, SARS-CoV2 recombinant protein S1/S2 added to a final concentration of 150 ng/μL, Biotin was added to 50 μM. After fully mixed, the reaction was proceeded on ice for 2 h. Biotinylaed proteins were desalted using Zeba™ Spin Desalting Columns 7K MWCO (Cat#89882, Thermo Fisher) according to the manufacturer’s instruction. Then we confirmed the biotinylation of S1/S2 using dot blotting assay.

### Surface staining and Flow Cytometry

Cryopreserved PBMCs were thawed and rested in 10 ml complete RPMI 1640 with 10% human AB serum (Gemini Bioproducts) at 37 °C for 3 h. PBMCs were washed with FACS buffer (PBS plus 2% FBS, Gibco), and then Fc blocking reagent (Meltenyi Biotec) was added for 15 min at room temperature, followed by three time wash with FACS buffer. Cells were then incubated for 30 min on ice with corresponding antibodies or Biotinylaed S1/S2 proteins. Antibodies used in the T cell and B cells surface marker staining are listed in Table S5 and S6. Then washed three times with FACs and acquired by FACS Verse (BD Biosciences, San Jose, CA). Data were analyzed by FlowJo software (Version 10.0.8, Tree Star Inc., Ashland).

### Intracellular cytokine staining and Flow Cytometry

PBMCs in wells of a 96-well plate with 100 μL complete RPMI 1640 and 10% human AB serum (Gemini Bioproducts) and Pen-Strep, were incubated 44 hours with 1uM of recombinant proteins (S1, S2 and nucleoprotein, N). Golgi-Plug containing Golgi-stop (BD Biosciences, San Diego, CA) and DNAase (Sigma, USA) with or without 1µg/mL Ionomycin plus 50ng/mL phorbol 12-myristate 13-acetate (PMA) were added another 4 hours into the culture. A stimulation with an equal amount of PBS was used as negative control. PMA plus ionomycin were included as positive controls. Cells were then washed with FACS buffer and surface stained for 30 minutes on ice, then fixed and permeabalized with BD Cytofix/Cytoperm solution (BD, Cat#554722) for 30 min at 4°C, followed by washing with perm/wash buffer (BD) and intracellular stained for 30 minutes on ice. Antibodies used in this assay were listed in Table S7. The gates applied for the identification of Perforin, GZMB and IFNγ positive cells were defined according the cells cultured with PBS for each sample. All samples were acquired on a BD FACSymphony cell sorter (BD Biosciences, San Diego, CA).

### Quantification and Statistical analysis

FlowJo 10 and GraphPad Prism 8.0.2 are used for data and statistical analyses. The statistical detail information of the experiments was provided in the respective Figureure legends. Correlation analyses were performed using Spearman and Mann-Whitney tests were applied for unpaired comparisons. One-way analysis was performed for more than two-group analysis. *P* values less than 0.05 were considered to be significantly statistically.

## Supplementary

**Figure. S1.**
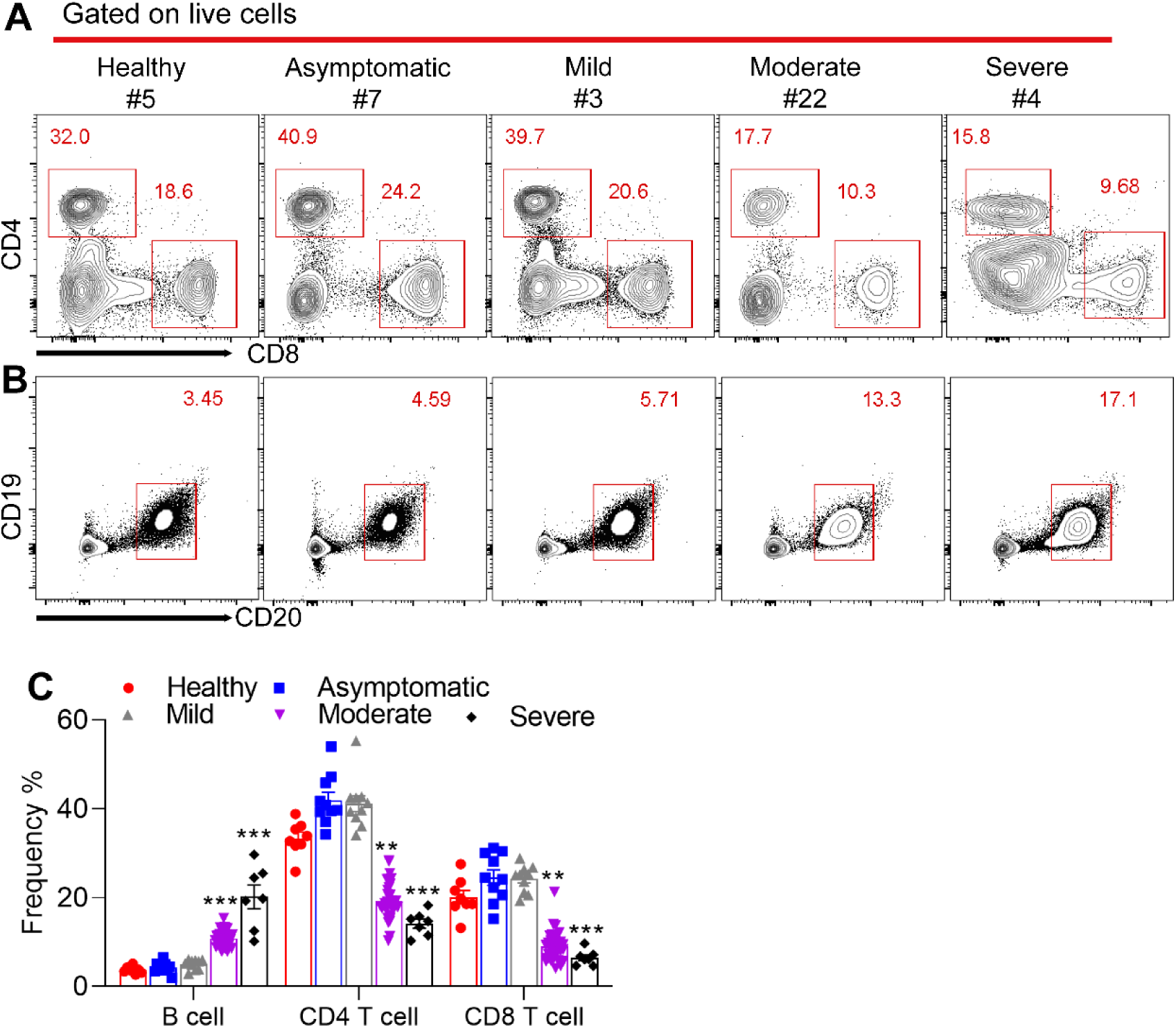
Frequency of B cells, CD4^+^T cells and CD8^+^T cells in total live PBMCs in COVID-19 patients with different severity. Samples of **a** and **b** were from Healthy (#5), Asymptomatic (#7), Mild (#3), Moderate (#22), severe (#4). **a, b**, Representative flow cytometry plots of CD4^+^ and CD8^+^ T cells (**a**) and B cells (**b**) in total live PBMCs in convalescent-phase COVID-19 patients. **c**, Percentages of CD4^+^ T cells, CD8^+^ T cells an B cells, summarized from (**a, b**), in COVID-19 recovered patients with different disease severity. Bars represent the mean ± SEM. *P* values were calculated based on Bonferroni of one-way analysis. ***, *p*<0.001, and **, *p*<0.01.

**Figure. S2.**
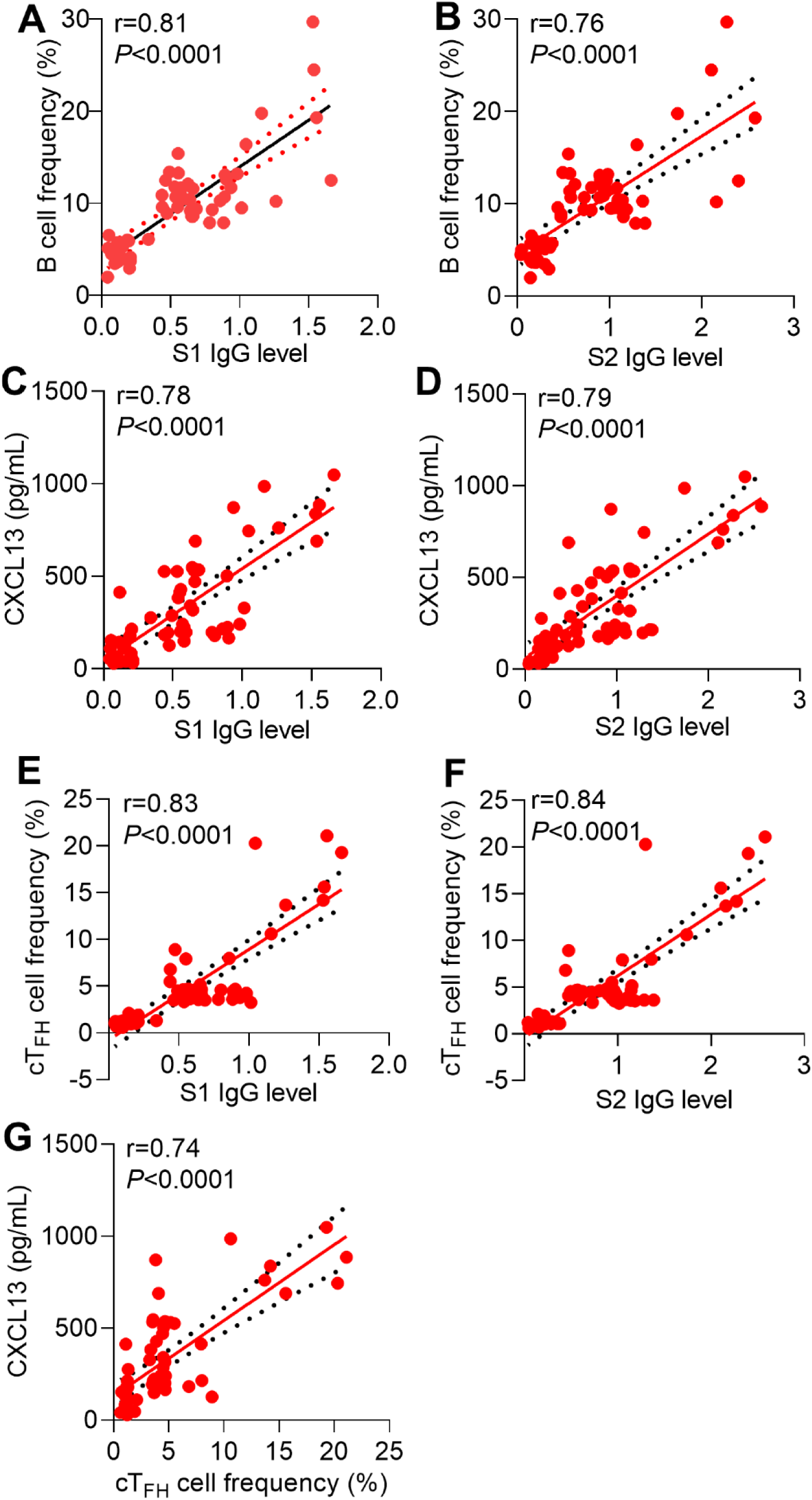
Correlations between B cells frequency, serum CXCL13 concentration, cTFH cells frequency and S1- or S2- specific IgG levels. **a, b**, Correlations between B cells frequency and S1- (**a**) or S2- (**b**) specific IgG levels. **c, d**, Correlations between serum CXCL13 concentration and S1- (**c**) or S2- (**d**) specific IgG levels. **e-f**, Correlations between cT_FH_ cells frequency and S1- (**e**) or S2- (**f**) specific IgG levels. **g**, Correlations between cT_FH_ cells frequency and serum CXCL13 concentration. Statistical comparisons were performed using Spearman correlation analysis.

**Table S1.** Characteristics of patients in this study.

|  | All<br>(n=67) | Healthy<br>(n=8) | Asymptomatic<br>(n=10) | Mild<br>(n=10) | Moderate<br>(n=32) | Severe<br>(n=7) |
| --- | --- | --- | --- | --- | --- | --- |
| **Age (years,**<br>**median,**<br>**IOQ)** | 67<br>(19-74) | 30.5<br>(25-38) | 21<br>(19-56) | 30<br>(19-56) | 39.5<br>(20-67) | 65<br>(43-74) |
| **Gender** |  |  |  |  |  |  |
| **Female** | 34 | 4 | 4 | 6 | 17 | 3 |
| **Male** | 33 | 4 | 6 | 4 | 15 | 4 |
| **Onset of**<br>**discharge to**<br>**sample**<br>**collection**<br>**(days,**<br>**median,**<br>**IOQ)** | 14<br>(10-17) |  | 13.5<br>(10-15) | 13.5<br>(10-14) | 13.5<br>(10-16) | 14<br>(14-17) |
| **Duration of**<br>**hospitalization**<br>**n** | 19<br>(7-51) |  | 24<br>(12-51) | 22<br>(12-31) | 16.5<br>(7-29) | 20<br>(13-24) |

1. Continuous variables were shown as the median (interquartile range, IQR). 2. All of 59 COVID-19 patients were tested positive qPCR for SARS-CoV-2 virus RNA upon hospital admission. Patients were diagnosed as severe when meeting any one of the following conditions: 1) anhelation ( $RR \geq 30/\text{min}$ ), 2) $SpO_2 \leq 93\%$ , 3) $PaO_2/FiO_2 \leq 300 \text{ mmHg}$ and 4) imageological diagnosis of significant progress ( $> 50\%$ ) in 24-48 hours. The moderate patients were diagnosed with respiratory symptoms, fever and imageological evidence of pneumonia. The mild patients were diagnosed with mild clinical symptoms and no imageological evidence of pneumonia. The asymptomatic patients were those without any clinical symptoms.

**Table S2.**
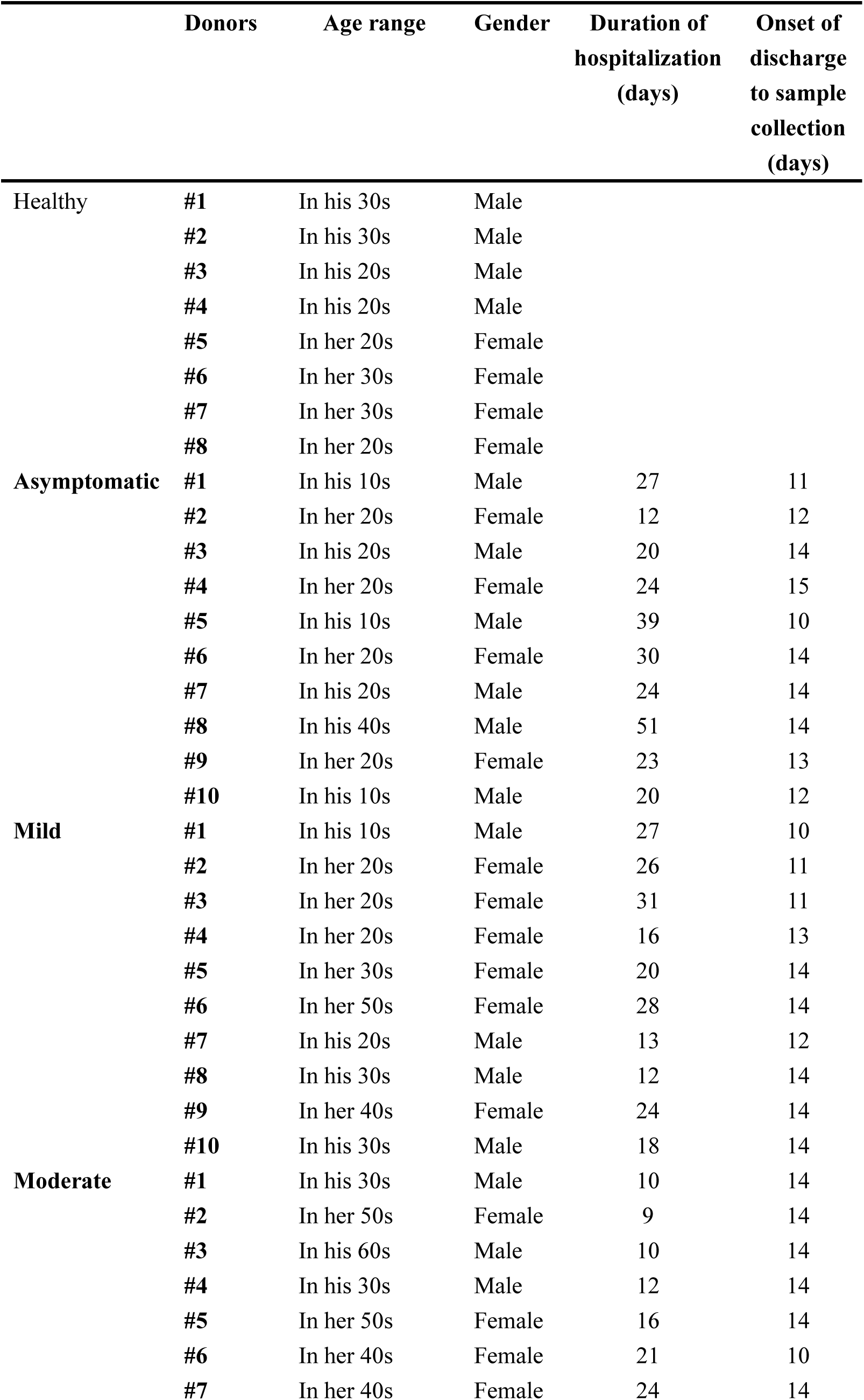
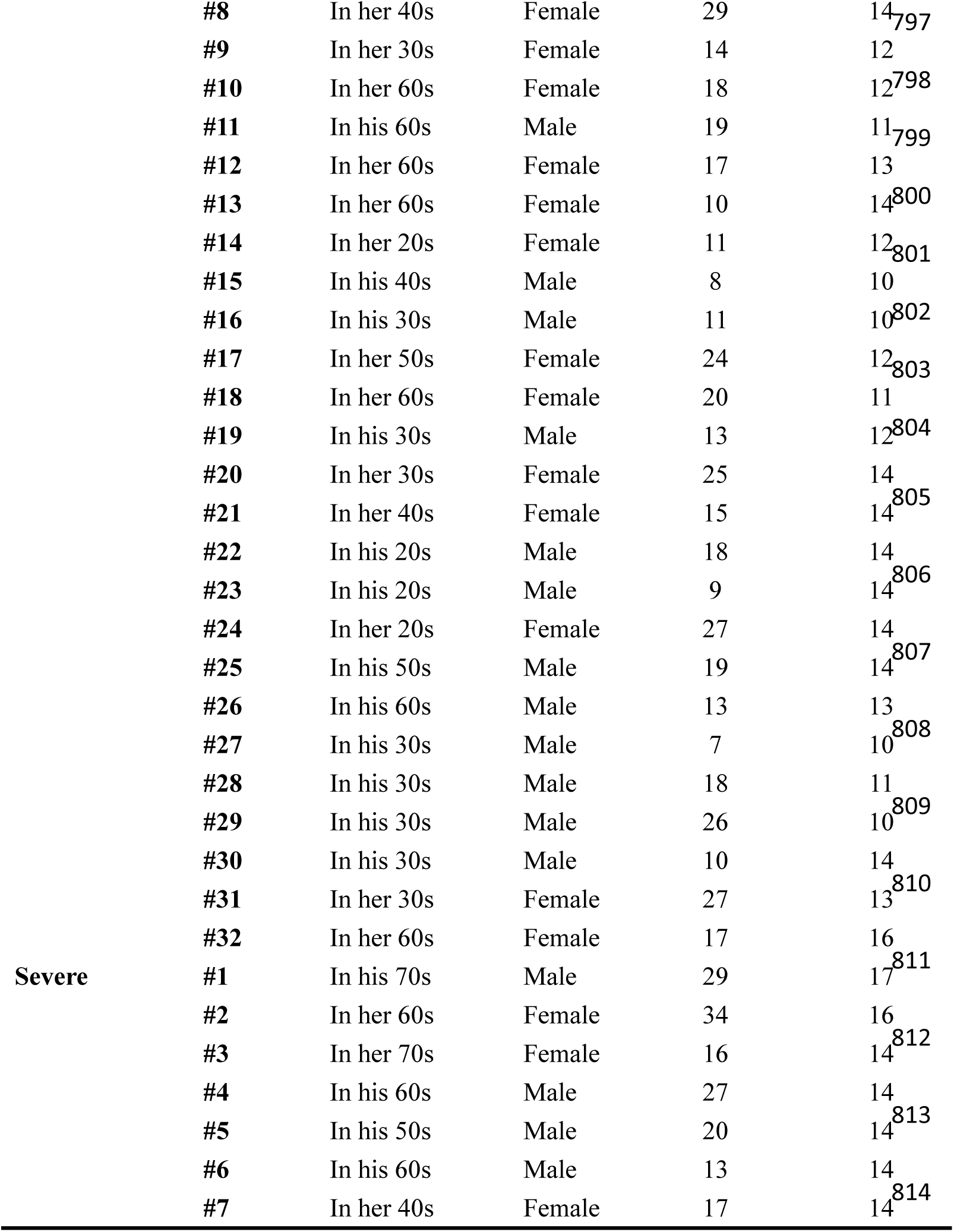
Characteristics of individual patients in this study.

**Table S3.**
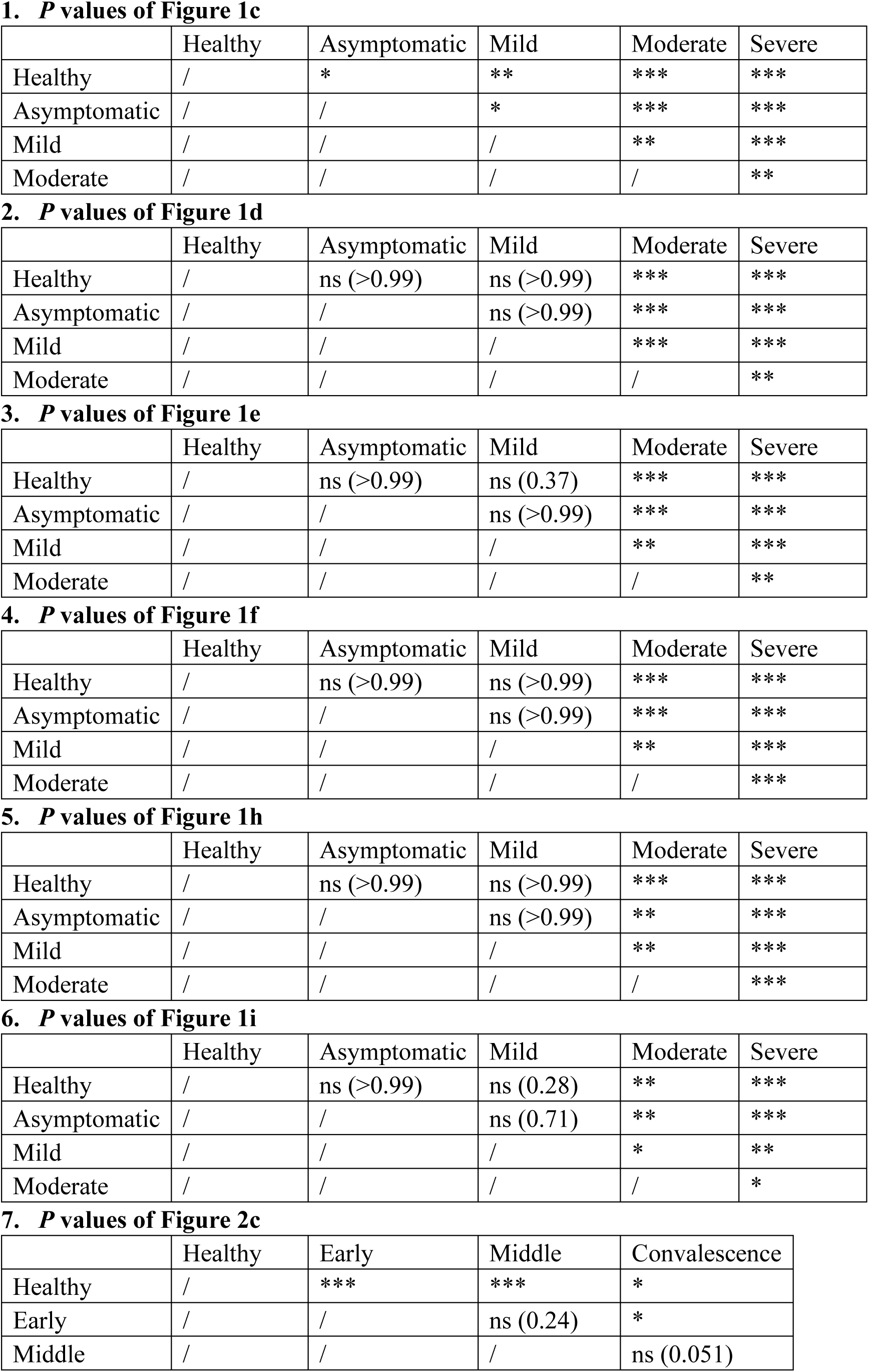
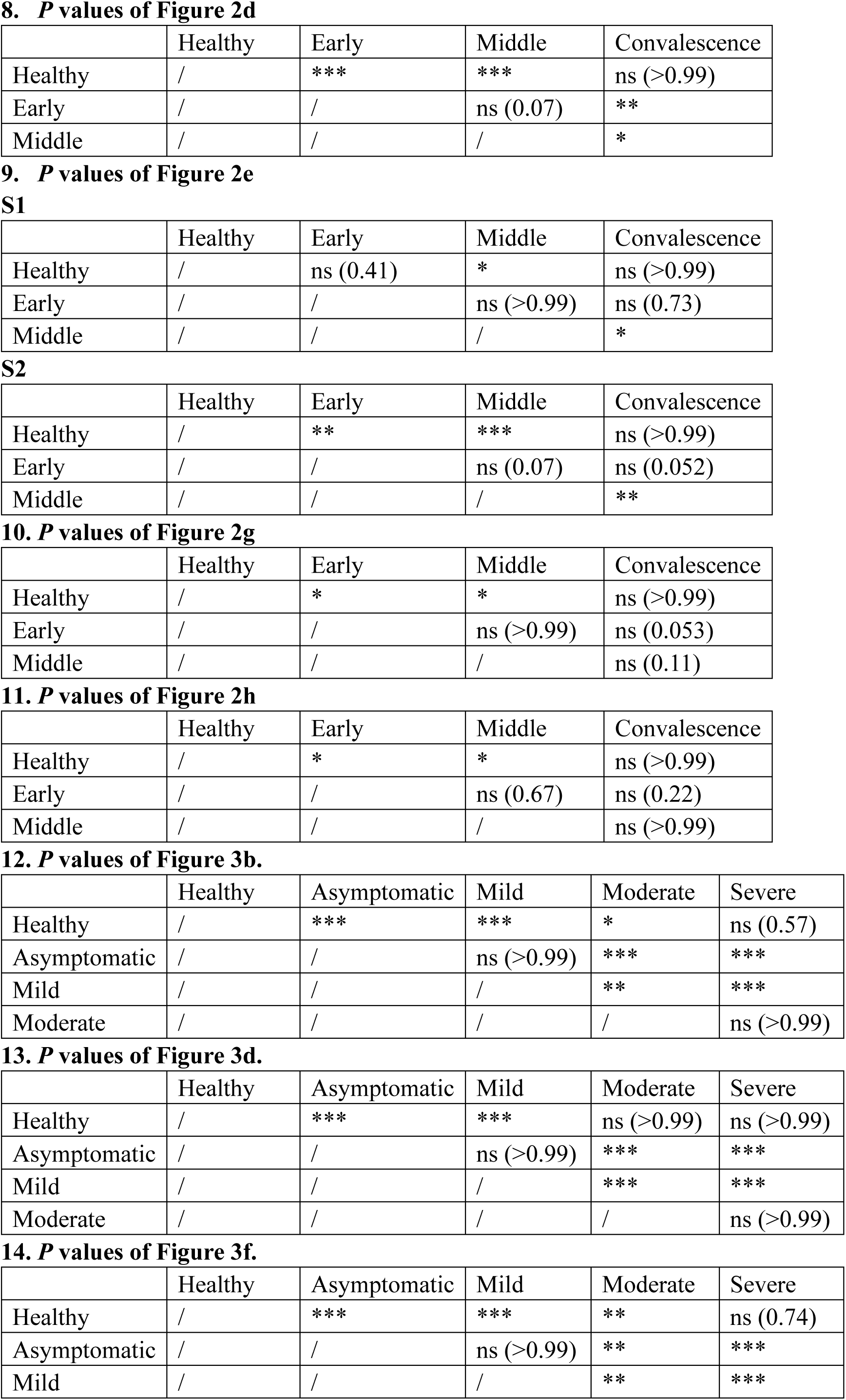
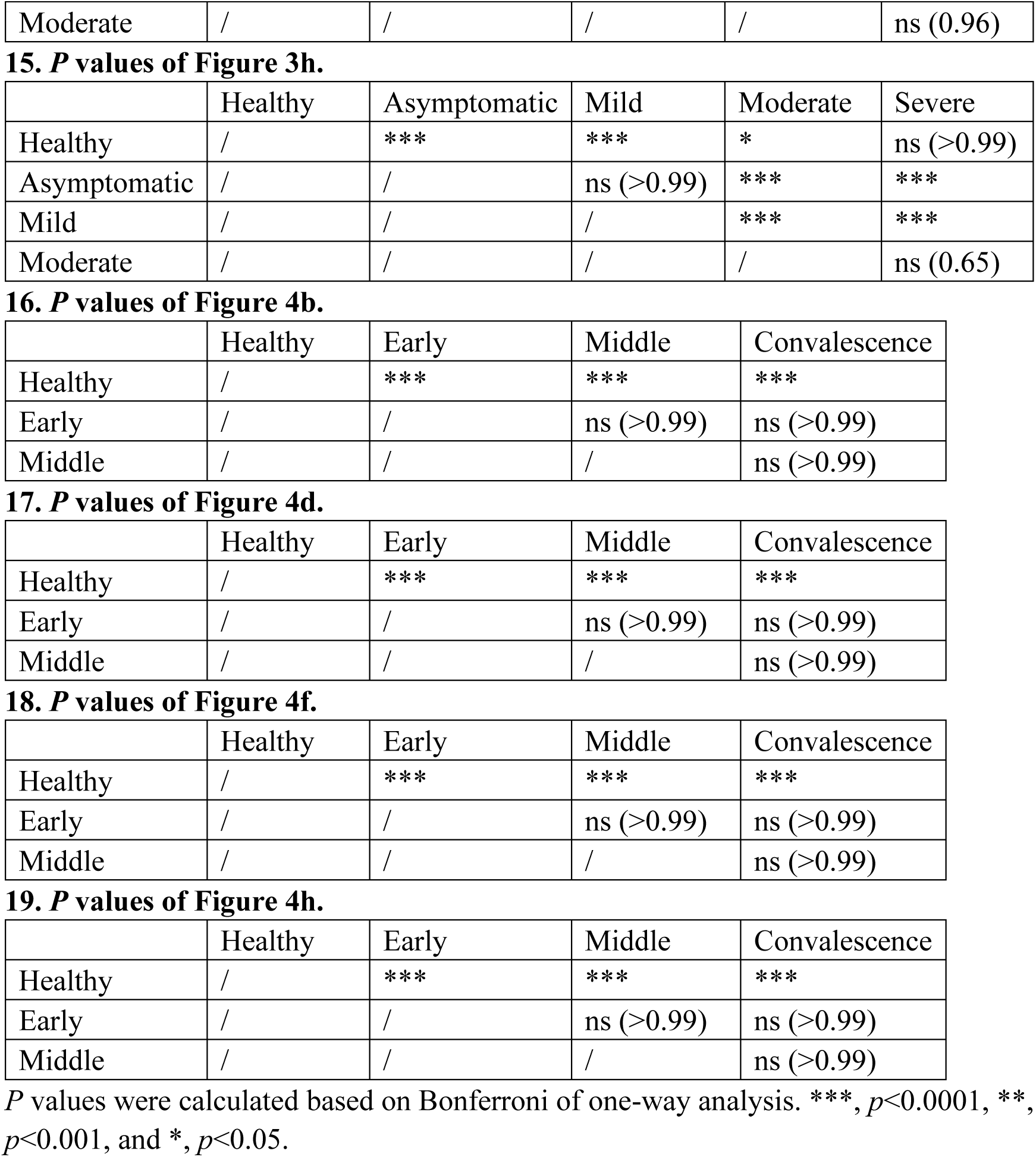
Summary of *P* values in Figure1–4.

**Table S4.**
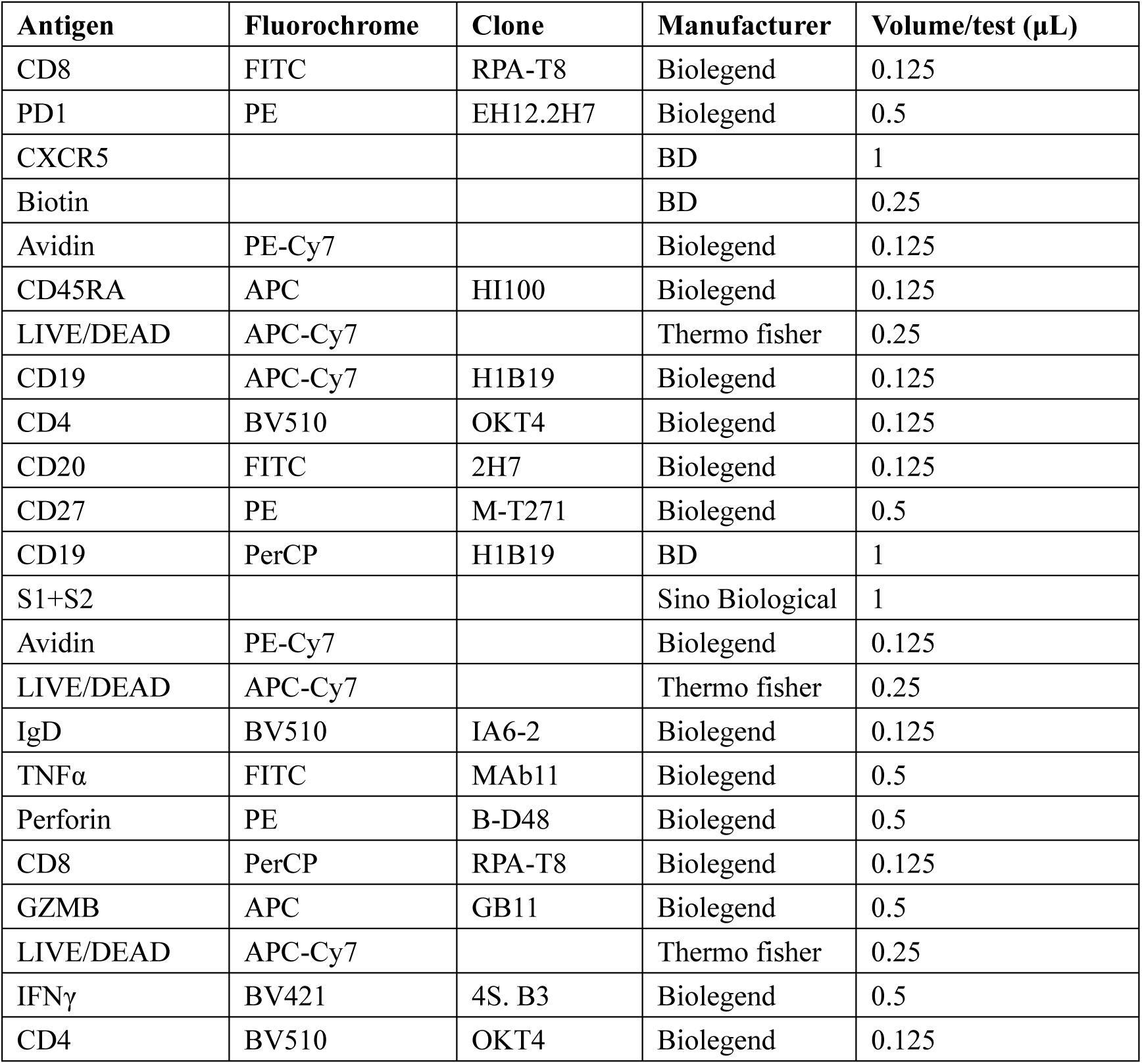
Flow cytometry antibodies in this study.

## Notes

### Competing Interest Statement

The authors have declared no competing interest.

### Author Declarations

The study received IRB approvals at Chongqing Public Health Medical Center (2020-023-01-KY).

